# COVID-19 is associated with relative ADAMTS13 deficiency and VWF multimer formation resembling TTP

**DOI:** 10.1101/2020.08.23.20177824

**Authors:** Adrian Doevelaar, Martin Bachmann, Bodo Hölzer, Felix S. Seibert, Benjamin S. Rohn, Frederic Bauer, Oliver Witzke, Ulf Dittmer, Michael Bachmann, Serap Yilmaz, Rita Dittmer, Sonja Schneppenheim, Nina Babel, Ulrich Budde, Timm H. Westhoff

**Author notes:** Author for correspondence Univ.-Prof. Dr. Timm H. Westhoff University Hospital Marien Hospital Heme Ruhr-University Bochum Medical Dept. I Hölkeskampring 40 44625 Herne Germany. authors contributed equally to the work (Adrian Doevelaar, Martin Bachmann). authors contributed equally to the work (Ulrich Budde, Timm H. Westhoff).

## Abstract

**Background:** Thrombotic microangiopathy (TMA) has been repeatedly described in COVID-19 and may contribute to SARS-CoV-2 associated hypercoagulability. The underlying mechanisms remain elusive. We hypothesized that endothelial damage may lead to substantially increased concentrations of Von Willebrand Factor (VWF) with subsequent relative deficiency of ADAMTS13.

**Methods:** A prospective controlled trial was performed on 75 patients with COVID-19 of mild to critical severity and 10 healthy controls. VWF antigen (VWF:Ag), ADAMTS13 and VWF multimer formation were analyzed in a German hemostaseologic laboratory.

**Results:** VWF:Ag was 4.8 times higher in COVID-19 patients compared to healthy controls (p< 0.0001), whereas ADAMTS13 activities were not significantly different (p = 0.24). The ADAMTS13/VWF:Ag ratio was significantly lower in COVID-19 than in the control group (24.4±20.5 vs. 79.7±33.2, p< 0.0001). Fourteen patients (18.7%) undercut a critical ratio of 10 as described in thrombotic thrombocytopenic purpura (TTP). Gel analysis of multimers resembled the TTP constellation with loss of the largest multimers in 75% and a smeary triplet pattern in 39% of the patients. The ADAMTS13/VWF:Ag ratio decreased continuously from mild to critical disease (ANOVA p = 0.026). Moreover, it differed significantly between surviving patients and those who died from COVID-19 (p = 0.001) yielding an AUC of 0.232 in ROC curve analysis.

**Conclusion:** COVID-19 is associated with a substantial increase in VWF levels, which can exceed the ADAMTS13 processing capacity resulting in the formation of large VWF multimers identical to TTP. The ADAMTS13/VWF:Ag ratio is an independent predictor of severity of disease and mortality. These findings render further support to perform studies on the use of plasma exchange in COVID-19 and to include VWF and ADAMTS13 in the diagnostic workup.

## Introduction

Coronavirus disease 2019 (COVID-19) is associated with hypercoagulation affecting both the venous and arterial vasculature. Venous thomboembolism occurs in up to one-third of patients in intensive care.^1^ Arterial thrombembolism events, such as stroke, limb ischemia, and myocardial infarction have been described in COVID-19 as well.^2,3^ Beyond these macrovascular events, autopsy studies have demonstrated microvascular thrombosis in the lungs.^4,5^ The pathogenesis of SARS Coronavirus 2 (SARS-CoV-2) induced hypercoagulation is incompletely understood. On one hand, endothelial injury is supposed to play a central role,^6^ on the other blood hyperviscosity and an imbalance of pro- and antithrombotic factors have been reported including elevated factor VIII, fibrinogen, and von Willebrand factor (VWF).^7^–^9^ Of note, there is an increasing number of reports on thrombotic events despite therapeutic plasmatic anticoagulation.

In some of our patients with COVID-19 we observed laboratory findings of microangiopathic hemolysis. Simultaneously, first cases of thrombotic microangiopathy (TMA) were reported by other groups in COVID-19 patients.^10,11^ To date, the pathophysiology of TMA in COVID-19 remains elusive. Based on the mechanisms underlying TMA in general, complement-mediation and a deficiency in ADAMTS13 may be considered. We investigated, whether the latter mechanism may play a pivotal role in this context.

As a blood glycoprotein involved in hemostasis Von Willebrand factor (VWF) is produced by endothelial cells and megakaryocytes.^12^ Endothelial cell activation is associated with the elevation of VWF. Accordingly, an increasing number of reports describe increased concentrations of VWF in COVID-19.^13,14^ The plasma protease ADAMTS13, a zinc-containing metalloprotease cleaves the large string-like molecules of VWF and is necessary to avoid accumulation of VWF multimers in the blood stream. In patients with ADAMTS13 deficiency accumulated VWF multimers cause thrombotic thrombocytopenic purpura (TTP). So far, ADAMTS13 has rarely been investigated in the context of COVID-19 and, in contrast to TTP, was normal or only mildly reduced.^11,15,16^ We therefore hypothesized that a massive elevation of VWF with clinically relevant VWF multimer abnormalities in COVID-19 patients may exceed the enzymatic capacity of ADAMTS13, which could contribute to the SARS-CoV-2 associated hypercoagulation.

To test this, we performed a prospective controlled trial on VWF antigen (VWF:Ag), ADAMTS13 activity and VWF multimer formation in a cohort of 75 COVID-19 patients. 10 healthy subjects served as control.

## Methods

We enrolled 75 patients, who had tested positive for SARS-CoV-2 by RT-PCR analysis of respiratory specimens (nasopharyngeal swab test or bronchoalveolar lavage). Patients were recruited at Ruhr-University Bochum, University of Duisburg-Essen, and Asklepios Klinikum Hamburg Harburg, Germany. The severity of COVID-19 ranged from mild to critical and was categorized according to the guidelines of the Robert Koch Institute, Germany. Patients with moderate and severe COVID-19 were recruited after the first symptoms occurred and a positive SARS-CoV-2 PCR result was available. For patients with critical disease, the recruitment took place at the intensive care unit, being diagnosed with COVID-19 in median 14 days before. The study was approved by the ethical committees of Ruhr-University Bochum (20–6886), University Hospital Essen (20–9214-BO) and the Medical Association Hamburg. Demographic and clinical characteristics of patients are summarized in Table 1.

**Table 1:** Clinical characterization of the patients with COVID-19

| Parameter | Study population with COVID-19 (n=75) |
| --- | --- |
| Age (years) | 66±16 |
| Female | 38 (50.7%) |
| Male | 37 (49.3%) |
| *Severity of COVID-19* |  |
| - mild | 3 (4%) |
| - moderate | 26 (34.7%) |
| - severe | 28 (37.3%) |
| - critical | 18 (24%) |
| *Laboratory findings* |  |
| - serum creatinine concentration (mg/dl) | 1.15±0.78 |
| - Glomerular filtration rate (ml/min, MDRD) | > 60 |
| - C-reactive protein (mg/dl) | 9.0±7.6 |
| - Lactate dehydrogenase (U/l) | 356.6±157.4 |
| - Hemoglobin (g/l) | 11.6±2.5 |
| - White blood cell count (/nl) | 8.3±5.3 |
| - Platelet count (/nl) | 257.8±182.7 |

### Measurement of ADAMTS13 activity, VWF antigen and VWF multimer analysis

ADAMTS13 activity (%) was analyzed from citrate-plasma using Technozym ADAMTS13 ELISA (Technoclone, Vienna, Austria).^17^ VWF:Ag (iU/ml) was measured using a sandwich ELISA with polyclonal antibodies.^18^ VWF multimer analysis was performed via sodium dodecyl sulfate agarose gel electrophoresis including a control sample containing normal VWF in each run to ensure proper conditions of the separation and blotting apparatus.^19^ The ADAMTS13/VWF:Ag ratio was calculated as (ADAMTS13 (IU/ml)/VWF:Ag (IU/ml) x 100). A ratio below 10 reflects the clinical situation of patients with acquired thrombotic thrombocytopenic purpura, where the vast majority of patients show ADAMTS13 activities below 10%. It is well known that a residual activity above 10% is sufficient to cleave the potentially harmful ultralarge VWF multimers. In the clinical situation of C0VID19 patients, the imbalance between the ADAMTS13 activity in the low reference range and the massively enhanced VWF leads to a TTP like situation, whenever the ADAMTS13/VWF ratio falls below 10.

### Statistics

Data are presented as mean ± standard deviation. Baseline differences in severity groups were compared by chi-squared tests for dichotomic parameters and by unpaired two-tailed t-tests for continuous parameters. Differences in ADAMTS13 activity, VWF:Ag, and ADAMTS13/VWF:Ag ratio between patients with COVID-19 and healthy controls and between surviving patients and patients, who ultimately died from COVID-19, were investigated by unpaired two-tailed t-tests. Receiver operating characteristic curves (ROC) were built to assess the predictive value for death from COVID-19. Univariate linear regression analysis was performed to investigate the association of age and laboratory findings on the ADAMTS13/VWF:Ag ratio in COVID-19 patients. P< 0.05 was regarded significant. Statistical analyses were performed using SPSS Statistics 25 (IBM, Chicago, USA) and Prism 8 (Graph Pad, San Diego, USA).

## Results

We enrolled 75 patients with SARS-CoV-2 infection and 10 healthy controls in the study. Measurements of VWF:Ag and ADAMTS13 were conducted successfully in all the patients. Mean age of the COVID-19 population was 66±16 years. Gender distribution was homogeneous with n = 38 being female (50.7%). Three patients (4%) had mild, 26 (34.7%) moderate, 28 (37.3%) severe, and 18 (24%) critical disease. Thirteen patients (17.3%) died. The control population had a mean age of 41±19 and was predominantly female. Table 1 summarizes demographic and clinical characteristics of the study population.

Table 2 presents the coagulation parameters of the study population and the control group. The VWF:Ag was substantially increased compared to the control group (4.03±2.18 IU/ml vs. 0.84±0.27 IU/ml, p < 0.0001, Figure 1A). ANOVA analysis showed no significant difference between the degrees of severity. ADAMTS13 activity was comparable to healthy controls (67.8±22.4% vs. 59.3±8.6%, p = 0.24; Figure 1B). There was a significant difference in ADAMTS13 activities, however, among the different severity degrees of COVID 19 (ANOVA p = 0.001; Figure 2B). The ratio of ADAMTS13/VWF:Ag was substantially lower in COVID-19 patients than in healthy controls (24.4±20.5 vs. 79.7±33.2, p<0.0001, Figure 1C). The ratio of ADAMTS13/VWF:Ag decreased continuously with the degree of COVID-19 severity (ANOVA p = 0.026; Figure 2C).

**Table 2:** Coagulation parameters in COVID-19 patients and healthy controls

| **Parameter** | **COVID-19**<br>**population (n=75)** | **Healthy controls**<br>**(n=10)** | **P** |
| --- | --- | --- | --- |
| D-dimer (mg/l) | 3.35±4.23 | 0.30±0.23 | < 0.0001 |
| Fibrinogen (mg/dl) | 536±142 | 361±60 | < 0.0001 |
| Partial<br>thromboplastin time<br>(s) | 32.0±13.2 | 29.3±3.3 | 0.494 |
| International<br>normalized ratio<br>(INR) | 1.17±0.37 | 0.95±0.07 | 0.001 |
| VWF:Ag (IU/ml) | 4.03±2.18 | 0.84±0.27 | < 0.0001 |
| ADAMTS13 (%) | 67.8±22.4 | 59.3±8.6 | 0.239 |
| ADAMTS13/VWF:Ag | 24.4±20.5 | 79.7±33.2 | < 0.0001 |

**Figure 1:**
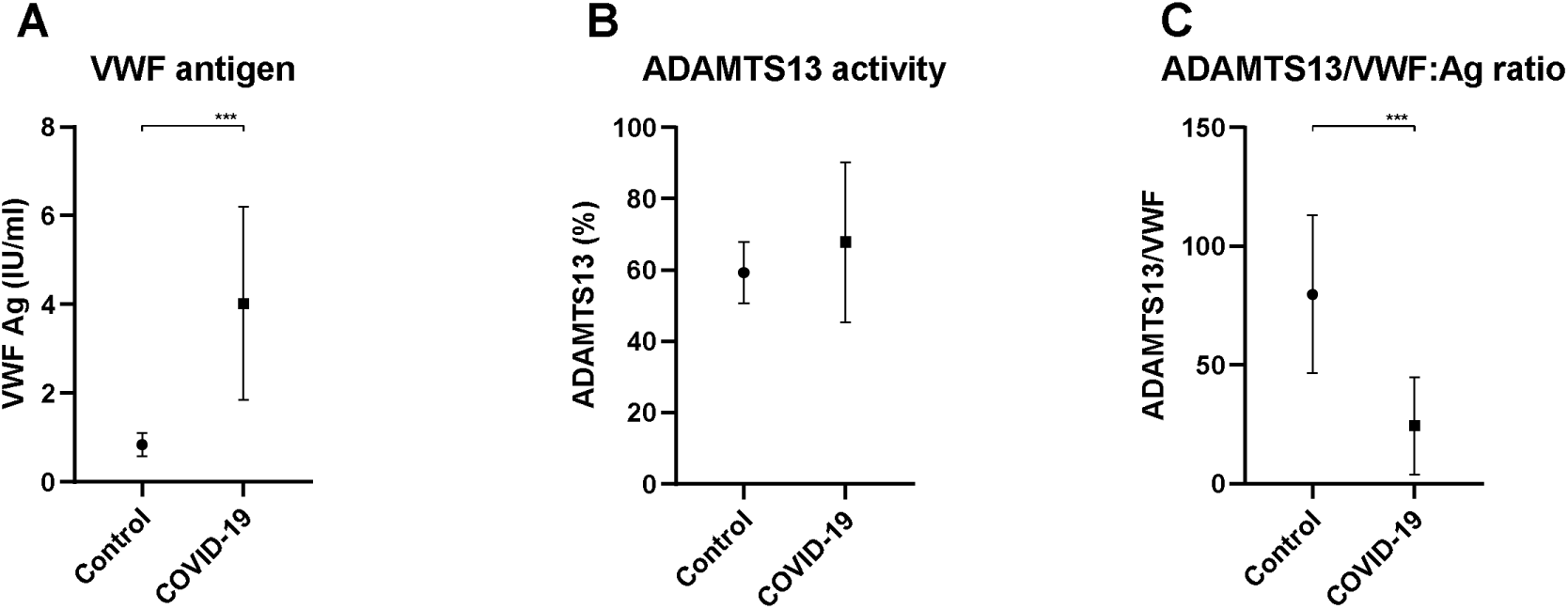
Von Willebrand Factor antigen (VWF:Ag), ADAMTS13 activity and the ratio of ADAMTS13/VWF:Ag in the population infected by SARS-CoV2 (n = 75) and healthy controls (n = 10).

**Figure 2:**
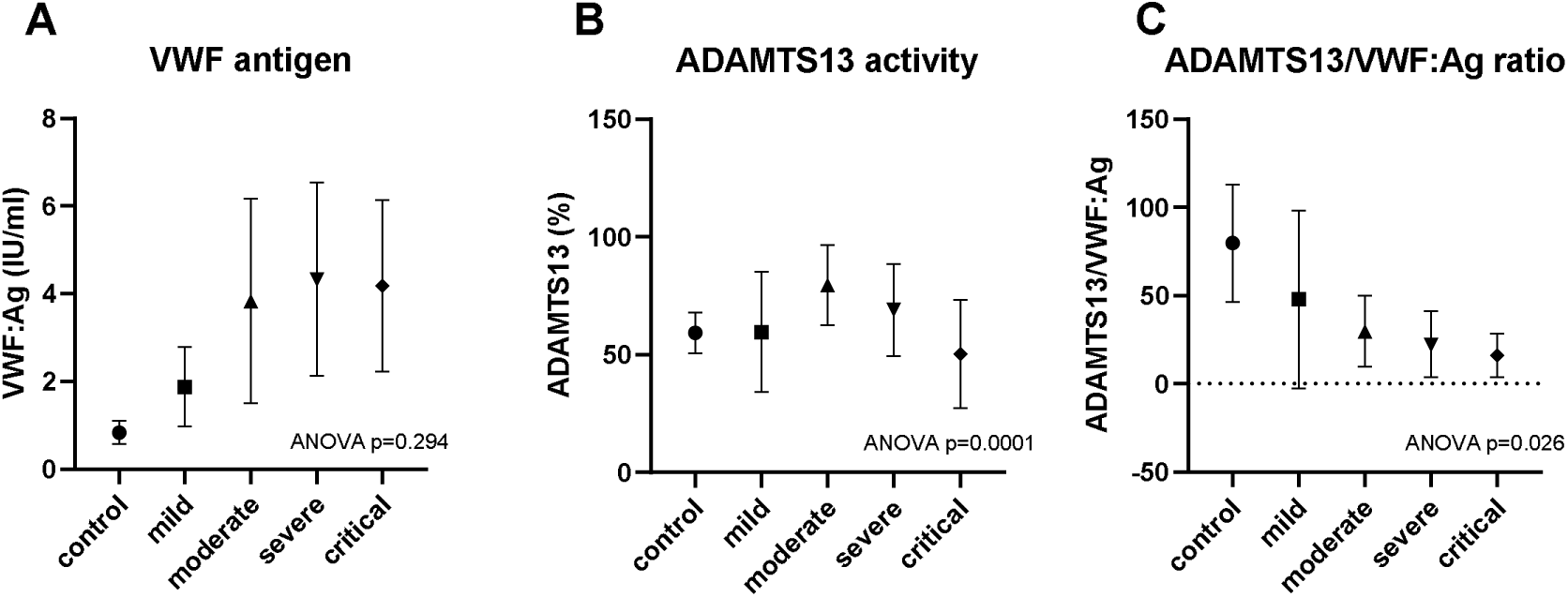
Von Willebrand Factor antigen (VWF:Ag), ADAMTS13 activity and the ratio of ADAMTS13/VWF:Ag in dependence of COVID-19 severity compared to healthy individuals. ANOVA was used to test for significant differences in patients with COVID-19. P< 0.05 was regarded significant.

Comparing patients who died from COVID-19 with those who survived, ADAMTS13 and ADAMTS13/VWF:Ag ratio were significantly lower in subjects who did not survive COVID-19 (72.6±20.4% vs. 45.2±18.0%, p< 0.001 for ADAMTS13 and 26.8±21.4 vs. 13.0±10.3, p = 0.001 for ADAMTS13/VWF ratio). VWF:Ag did not significantly differ between survivors and those who died (p = 0.181). ROC analyses for ADAMTS13/VWF:Ag and death from COVID-19 provided an area under the curve (AUC) of 0.232. The results are presented in Figure 3.

**Figure 3:**
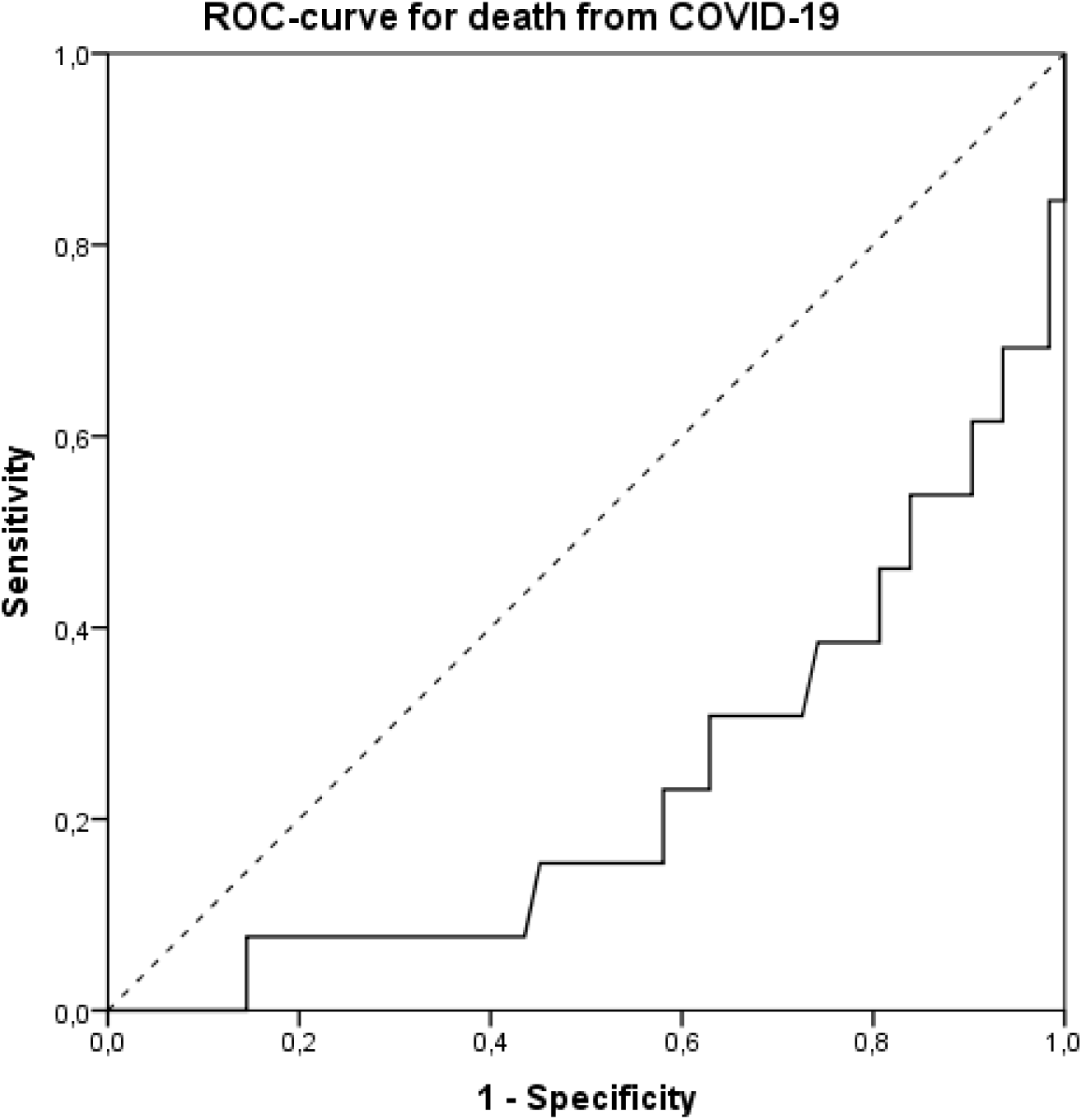
ROC analysis for death from COVID-19 in dependence of the ratio of ADAMTS13/VWF:Ag.

Univariate linear regression analysis was performed to investigate the association of age and laboratory findings on the ADAMTS13/VWF:Ag ratio in COVID-19 patients (Table 3). Age (T-3.269, p = 0.002), C-reactive protein (T −2.777, p = 0.007), platelet count (T 2.345, p = 0.022), hemoglobin (T 2.401, p = 0.019) and partial thromboplastin time (T −2.576, p = 0.012) revealed significant associations with the ADAMTS13/VWF:Ag ratio. In multivariate linear regression for these parameters, a significant association remained for age (p = 0.005), hemoglobin (p = 0.031) and platelet count (p = 0.017).

**Table 3:** Univariate linear regression analysis using ADAMTS13/VWF:Ag as dependent variable in the COVID-19 population

|  | **Regression**<br>**coefficient**<br>**(B)** | **Standard**<br>**error** | **Beta** | **T** | **P** |
| --- | --- | --- | --- | --- | --- |
| Age | -0.457 | 0.140 | -0.357 | -3.269 | **0.002** |
| Serum<br>creatinine | -5.194 | 3.058 | -0.196 | -1.699 | 0.094 |
| Hemoglobin | 2.251 | 0.937 | 0.271 | 2.401 | **0.019** |
| White blood<br>cell count | -0.193 | 0.455 | -0.050 | -0.424 | 0.673 |
| Platelet count | 0.03 | 0.013 | 0.265 | 2.345 | **0.022** |
| C-reactive<br>protein | -0.841 | 0.303 | -0.311 | -2.777 | **0.007** |
| INR | -12.468 | 6.505 | -0.220 | -1.917 | 0.059 |
| Partial<br>thromboplastin<br>time | -0.455 | 0.177 | -0.290 | -2.576 | **0.012** |
| Lactate<br>dehydrogenase | 0.000 | 0.016 | 0.002 | 0.013 | 0.989 |

VWF multimers were analyzed by gel analysis and categorized as large (>10 oligomers), intermediate (6–10 oligomers), and small (1–5 oligomers). Gel analysis was successful in all but one patient. In TTP large and ultralarge multimers accumulate in the microthrombi and therefore show reduced concentrations in the circulation. 25% of the samples showed a normal amount of large VWF multimers (80–100% of the control sample), 50% showed a mild reduction (60–79% of the control sample), and in 25% they were severely reduced (< 60% of the control sample). Large multimers in COVID-19 patients were significantly lower than in healthy pool samples (68.69±16.16% vs. 124.20±8.89%, p< 0.0001). The Triplet structure of the small multimers showed smear – an indicator of ADAMTS13 dysfunction – in 39% of the patients. There was a loss of largest multimers of 13–29% in COVID-19 and 18–47% in TTP patients. Figure 4 presents a representative gel of a healthy control, three COVID-19 patients with severe disease, and three patients with TTP. It illustrates the similarity of plasmatic multimer composition in COVID-19 and TTP.

**Figure 4:**
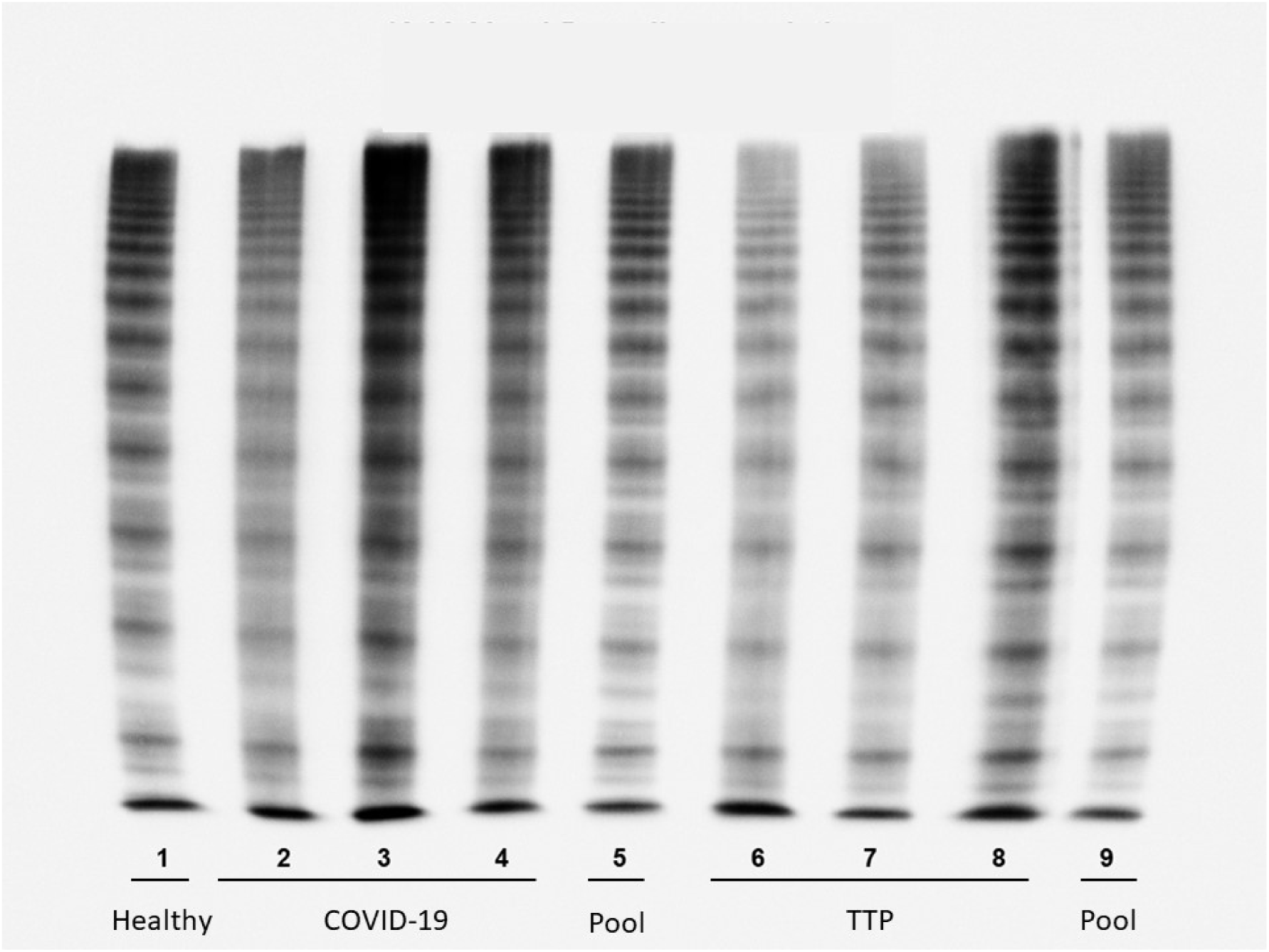
Von Willebrand factor multimers in a medium resolution gel (1.8% LGT-agarose) of (1) a normal person, (2–4) three patients with COVID-19, (5, 9) a normal plasma pool, and (6-8) three patients with acute TTP prior to initiation of treatment. The gel shows smear in the patient lanes of both COVID-19 and TTP. There was a loss of largest multimers of 13–29% in COVID-19 and 18–47% in TTP patients.

## Discussion

In TTP the ratio of ADAMTS13/VWF:Ag is decreased below a critical level of 10. In this situation the amount of ADAMTS13 does not suffice to avoid the generation of ultralarge VWF multimers. The present study shows that this scenario occurs in the context of COVID-19 as well. In TTP, the low ADAMTS13/VWF:Ag ratio results from an absolute deficiency of ADAMTS13 with activity levels < 10%. In the vast majority of TTP patients, ADAMTS13 deficiency is acquired, e. g. by antibody formation, while in a minority, it is inherited known as Upshaw–Schulman syndrome.^20,21^

We observed no single case of critical absolute deficiency of ADAMTS13. ADAMTS13 has rarely been measured in COVID-19 before.^11,15,16^ In line with our findings, these isolated publications do not report a critical deficiency of ADAMTS13 either. Whereas there is no absolute deficiency, however, the massive production of VWF exceeds the processing capacity of the protease leading to a relative deficiency of ADAMTS13. Thus, in 18.7% of the study population the ADAMTS13/VWF:Ag ratio fell below the critical threshold of 10.

Endothelial damage is a consistent finding in autopsy series of patients with COVID-19. It is supposedly mediated both by direct infection of endothelial cells, which express the ACE2 receptor, and secondary damage due to microvascular inflammation.^22,23^ Endothelial injury may thereby constitute a crucial common trait in the multiorgan manifestations of COVID-19. Endothelial activation is associated with an increased production of VWF, which likely explains the massive upregulation in the present study population. To avoid formation of ultralarge VWF multimers in states of endothelial damage with increased VWF generation, there is an evolutionary preserved large reserve of ADAMTS13. Under physiological conditions 10% of the available ADAMTS13 is sufficient to prevent formation of VWF multimer. Our present findings show, however, that the massive increase of VWF in COVID 19 exceeds the reserve of ADAMTS13. In this patient population VWF levels were elevated to a mean of 4.03 iU/ml and were thus 4.8 times higher than the average VWF levels in the healthy control group population. It may be speculated, that the ubiquitous endothelitis causes the extraordinarily large increase in VWF plasma levels. The ADAMTS13/VWF:Ag ratio is therefore a valuable biomarker to detect a critical relative deficiency of ADAMTS13.

Gel analyses support the hypothesis of an exceeded protease capacity of ADAMTS13: They showed significantly reduced concentrations of large VWF multimers compared to healthy subjects with almost 40% of the COVID-19 patients having the typical TTP pattern of smear in triplet structure analysis of the small multimers. Figure 4 illustrates the similarity in multimer composition of the two diseases despite the normal absolute concentrations of ADAMTS13.

A pathophysiological role of relative ADAMTS13 deficiency is further supported by the finding that the ADAMTS13/VWF:Ag ratio predicted mortality in the study cohort. With an AUC of 0.232 in ROC analysis the ratio demonstrated a quite high predictive value for fatal outcome. Interestingly, ADAMTS13/VWF:Ag continuously decreased with the level of COVID-19 severity. Hence, the ratio is not only a predictor of mortality but also reflects the intensity of morbidity. Moreover, a clinical relevance is supported by the significant association of the ADAMTS13/VWF:Ag ratio with platelets and hemoglobin in regression analysis. Both platelets and hemoglobin concentrations are reduced in TMA.

Based on the autopsy findings of multiple venous thromboses and thrombembolisms plasmatic anticoagulation was recommended for COVID-19 patients.^24^ In the Hamburg study population almost all patients who were treated in the ICU and died were autopsied. Interestingly, the patients had multiple thromboses and thrombembolisms even though they all had been treated with heparin or argatroban in therapeutic doses from the beginning of their stay on ICU. These findings support the hypothesis that there must be an additional pathogenic mechanism, which cannot be sufficiently addressed by plasmatic anticoagulation therapies.

The present study identifies massive VWF release with relative deficiency of ADAMTS13 as a candidate mechanism of TMA in COVID-19. Besides this mechanism, there is initial evidence that complement activation may contribute to TMA as well. Complement activation has been reported in mouse models of SARS.^25^ More importantly, histological studies of SARS-CoV-2 pneumonitis revealed septal capillary luminal fibrin accumulation and accumulations of terminal complement components C5b-9 and C4d in the microvessels, consistent with an activation of the alternative complement pathway.^4^ Attempts to address complement overactivation in COVID-19 with eculizumab are ongoing.

Our findings suggest two further therapeutic approaches. First, caplacizumab has recently been approved for acquired TTP. Caplacizumab is an anti-VWF nanobody that inhibits the interactions of ultralarge VWF multimers and platelets.^26^ Moreover, plasma exchange may be a promising therapeutic approach for patients with COVID-19 and the laboratory constellation of TMA. Plasma exchange eliminates excessive VWF, delivers ADAMTS13, and –additionally – is able to reduce complement activation. There are already first case series, in which plasma exchange was used in order to attenuate circulating cytokines and inflammatory mediators in critically ill patients with COVID-19.^27,28^ In three patients treated with plasma exchange, C-reactive protein and IL-6 levels decreased.^27^ In a cohort of 31 patients in Oman, 11 critically ill patients underwent plasma exchange. Plasma exchange was associated with higher extubation rates, and lower 14 days and 28 days all-cause mortality.^28^ In line with our findings, these reports provide a rationale for the conductance of prospective randomized trials, that are now under way.

Our study is limited by the cross-sectional assessment of VWF:Ag and ADAMTS13. Future studies should address the dynamics of VWF and ADAMTS13 during COVID-19. Moreover, the critical importance of the ADAMTS13/VWF:Ag ratio has to be confirmed in larger patient cohorts.

Without therapeutic interventions, the mortality of patients with TMA is high. The present study enlarges our understanding of the mechanisms underlying TMA in COVID-19. It appears reasonable to include VWF:Ag and ADAMTS13 in the diagnostic workup of COVID-19, at least in laboratory constellations of microangiopathic hemolysis. Until data from prospective clinical trials on the use of plasma exchange in COVID-19 are available, clinical decision for its use should be made in an individualized manner.

## Data Availability

No external datasets or supplementary material at other repositories.

## Author contributions

T.H.W. supervised the project and wrote the manuscript with support from A.D., U.B., M.B., N.B.and S.Y.

A.D., B.H., N.B., F.S.S, B.S.R., F.B., O.W. and U.D. contributed to data collection.

A.D., T.H.W. and U.B. analysed the data.

U.B., R.D. and S.S. performed the measurements on VWF and ADAMTS13.

All authors discussed the results and contributed to the final manuscript.

## Abbreviations

(Covid-19): Coronavirus disease 2019
(SARS-CoV-2): SARS Coronavirus 2
(VWF): Von Willebrand factor

## Acknowledgements

We thank the laboratory staff Kerstin Will, Claudia Fiedelschuster, Barbara Schocke and Dr. Antje Pieconka of MEDILYS, Hamburg, for their indefatigable efforts in this study in times of excessive routine workloads.

## Conflict of interest

The authors have declared that no conflict of interest exists.

## Source of funding

None.

## Notes

### Competing Interest Statement

The authors have declared no competing interest.

### Clinical Trial

Multi thematic study process ongoing

### Funding Statement

No external funding was received for this study.

### Author Declarations

The study was approved by the ethical committees of Ruhr-University Bochum (20-6886), University Hospital Essen (20-9214-BO) and the Medical Association Hamburg.

